# Characterizing the COVID-19 illness experience to inform the study of post-acute sequelae and recovery: a qualitative study

**DOI:** 10.1101/2021.03.10.21253330

**Authors:** Edda I. Santiago-Rodriguez, Andres Maiorana, Michael J. Peluso, Rebecca Hoh, Viva Tai, Emily A. Fehrman, Yanel Hernandez, Leonel Torres, Matthew A. Spinelli, Monica Gandhi, J. Daniel Kelly, Jeffrey N. Martin, Timothy J. Henrich, Steven G. Deeks, John A. Sauceda

## Abstract

We aimed to characterize the variability in the illness experience and recovery process from COVID-19. We conducted in-depth individual interviews with participants enrolled in the Long-term Immunological Impact of Novel Coronavirus (LIINC) cohort study in San Francisco, California from June through October of 2020. Participants were adults who had a previously confirmed positive SARV-CoV-2 nucleic acid amplification test result, had recovered or were recovering from acute infection, and underwent serial evaluations at our clinical research center. We purposefully sampled 24 English- and Spanish-speaking adults with asymptomatic, mild and severe symptomatic infection, including those who were hospitalized, and those with HIV co-infection. Half of our sample (50.0%) identified as Latinx/Hispanic and most of the participants were men (62.5%). We used thematic analysis to characterize the illness experience, recovery process, and mental health impact of experiencing COVID-19 and present clinical data for each participant. Emergent themes were: (1) across symptom profiles and severity, experiencing COVID-19 was associated with psychological distress, (2) among participants with symptomatic infection, the illness experience was characterized by uncertainty in terms of managing symptoms and recovery, and (3) despite wide-ranging illness experiences, participants shared many common characteristics, including health information-seeking behavior facilitated by access to medical care, and uncertainty regarding the course of their illness and recovery. COVID-19 was associated with elevated levels of psychological distress, regardless of symptoms.

## INTRODUCTION

There is an urgent need to understand the impact and optimal management of the variability in the disease experience and recovery from post-acute sequelae of SARS-CoV-2 infection (PASC).^1^ Our early understanding of the *lived experiences* and challenges with recovery from COVID-19 was mainly from reports in media outlets, ^2-5^ and editorials, letters, and case reports in scientific journals.^6-8^ While these reports and editorials highlight unique disease experiences and areas to prioritize for future research, they do not leverage data from personal experiences with COVID-19 through formal qualitative research. Such research methods can characterize the variability in how people define and experience their disease and recovery,^9^ which can offer insights to optimize management of persistent symptoms of COVID-19.

COVID-19 has produced wide-ranging physical, neurological and mental health effects, many of which are yet to be fully understood,^10,11^ leaving many individuals in a state of uncertainty. However, there are lessons that can be drawn from research on chronic disease management where prognoses are uncertain, such as the potential benefits of information-seeking behavior^12^ and leveraging a person’s healthcare empowerment,^13^ both of which describe how individuals process, understand, seek and act on disease information, especially for coping with uncertainty and chronic disease.^14,15^ In this regard, qualitative research methods are a useful approach to characterize the processing of the COVID-19 diagnosis, illness experience, and recovery process.^16^ Furthermore, characterizing the illness experience and recovery trajectory in persons with a history of COVID-19 may inform the management of post-acute sequelae given the uncertainties that remain regarding its pathogenesis and emerging evidence that not all individuals experience a rapid or complete recovery.

### Objective

We conducted in-depth interviews with a diverse group of people with previously documented SARS-CoV-2 infection who had entered the “recovery phase” from COVID-19 and were able to attend at least one in-person visit at our study center. The group included persons who had experienced a spectrum of severity of the acute SARS-CoV-2 illness, including those who had asymptomatic infection, mild disease, and severe disease requiring hospitalization. We also included people with co-infections, such as people living with HIV (PLWH). We were interested in examining the spectrum of lived experiences following COVID-19 diagnosis. Participants in this study were enrolled in a parent cohort study, the Long-term Immunological Impact of Novel Coronavirus (LIINC) COVID-19 recovery cohort study.^17^ LIINC explores immunologic responses at different stages of convalescence to understand the effects of SARS-CoV-2 over time, and is based within an academic care and research center in an infectious diseases division at the University of California, San Francisco (UCSF). The clinical course for each participant interviewed will be provided to contextualize the participant interview data from this qualitative study.

## METHODS

### Ethics approval

The University of California, San Francisco Committee on Human Research reviewed and approved the parent study (LIINC) and this qualitative study. All participants in the parent study signed a written informed consent and provided written consent to be referred to participate in this qualitative study. Participants provided verbal consent for the interview and written consent for review of their clinical data.

### Eligibility Criteria

The LIINC study is based within the UCSF Division of HIV, Infectious Diseases, and Global Medicine and the Division of Experimental Medicine at San Francisco General Hospital (SFGH). Beginning in April 2020, participants with documented SARS-CoV-2 infection confirmed on nucleic acid amplification testing were eligible to enroll during the early recovery phase from COVID-19 (defined as 3-10 weeks following illness onset). Any adult with confirmed infection was eligible; eligibility was not contingent on any particular clinical characteristics of acute infection or recovery. Participants were recruited through clinician and participant self-referral, an online web portal (www.liincstudy.org), clinicaltrials.gov (NCT04362150), local media advertisements, and recruitment via the electronic medical record system. Following enrollment, participants were monitored monthly for up to 4 months and quarterly thereafter. Assessments included detailed clinical questionnaires administered by a trained clinical research coordinator under the supervision of a study physician (including demographic information, characterization of the acute illness including the circumstances of diagnosis and treatment, identification of any symptoms during the acute and recovery phases, comorbid medical conditions and concomitant medications, substance use), and biospecimen collection (including blood, whole unstimulated saliva, and gingival crevice fluid collection). Participants from the LIINC study were then enrolled in this separate qualitative study if interested.

### Consent for the qualitative study and interview procedures

Clinical staff provided a general overview of the qualitative study to LIINC participants; willing participants were then referred to the social science research team who conducted this study. A total of 42 participants were referred by the LIINC staff. The social science research team contacted potential participants through email, phone call, or text message based upon their stated preferences. Participants who were reached and available to participate were scheduled for a semi-structured in-depth interview through privacy-compliant video conference software or telephone. We used a purposeful sampling method by having weekly discussions with the LIINC team during active enrollment about the demographic characteristics of their referrals based on ethnicity/race, HIV status, language and occupation. The purpose of this sampling strategy was to ensure that diverse COVID-19 experiences and participant demographic characteristics were captured.

Interviews were conducted between June and October 2020. Interviews were of 30 to 90 minutes in duration and were conducted in English or Spanish by two fully bilingual qualitative researchers. The semi-structured interview guide was designed to explore areas related to COVID-19 such as: knowledge, prevention methods, symptoms, testing experiences, different recovery experiences, access to resources, housing stability, food insecurity and economic stability. Interviews were audio-recorded and transcribed ad verbatim. After the interview, each participant received a $50 gift card. We interviewed until saturation was evident (redundancy in narratives, no new insights), which occurred after 24 interviews.

### Analysis Plan

We used thematic analysis^18,19^ to systematically identify and organize patterns of meanings throughout a set of interview transcripts. A thematic analysis serves as a flexible method for explorative studies like this where you don’t have a clear idea of what theme patterns you are searching for in the data. All interview transcripts were uploaded onto Dedoose, a web-based program to organize and manage qualitative data.^19^ Two authors (EIS and AM) then reviewed a subset of two transcripts to identify initial impressions and develop an initial codebook. The codebook was developed using deductive and inductive codes based on the interview questions and themes emerging from the interviews. Each transcript was coded by a bilingual primary coder (social and community psychologist) and reviewed by a bilingual secondary coder (medical anthropologist). For this analysis, we reviewed the coded texts in their original language related to symptoms, treatment and recovery experiences among participants (only English text and translations are provided in the results). The team then had periodic sessions approximately every other week for two months to discuss and summarize the texts. Emerging and preliminary results were discussed with the senior author (health psychologist) and one of the LIINC co-principal investigators (infectious disease physician and researcher).

We also summarized clinical evaluation data from participants’ initial LIINC visit. These data were anonymized to protect confidentiality and are presented below participants’ first quotes to help contextualize their narratives. We present self-reported demographic information, exposures, symptoms, symptom onset and severity, presence of persistent symptoms, and treatments administered for COVID-19, and mental health during their illness course. For asymptomatic infections, we report the date of the positive SARS-CoV-2 test result, whereas for symptomatic infection, we report date of symptom onset. The clinical evaluation also asked participants with <u>symptomatic infection</u> to volunteer to rate how good/bad their health during the worst point of illness with COVID-19, from 0 (worst) to 100 (best). We labeled this score their *health score*. For participants with <u>asymptomatic infection</u>, the *health score* represents how good/bad their self-perceived health was immediately before their positive test result. Not all participants chose to answer all clinical evaluation questions, which is denoted by the phrase “none reported.”

## RESULTS

### Demographics

Twenty-four individuals participated in the qualitative study. The median patient age was 49 and most of the participants were men (62.5%) and white (66.7%). Half of our sample (50.0%) identified as Latinx/Hispanic. Nine participants reported they were hospitalized due to COVID-19, six of whom required oxygen support. Out of the nine, three were transferred to the intensive care unit, and two were intubated. Eight participants were living with HIV. Six participants completed the interview in Spanish.

We started all interviews by asking about how participants learned about COVID-19 at the start of the pandemic and where they sourced their information. We then transitioned to discussing participants’ experiences with COVID-19, the emotional impact of having a confirmed COVID-19 diagnosis, and the factors influencing the recovery process. To protect confidentiality, we report participants by identification numbers and modified data (e.g., dates) to maximize confidentiality.

### Asymptomatic Infection

Importantly, although some participants were categorized as having asymptomatic infection, they described deep concern over their diagnosis and uncertainty with potential illness and highlighted the benefits of having support and access to medical care. Participant #8, whose health score was 65/100 before diagnosis, stated that his experience with COVID-19 was informed by living with HIV and other comorbidities. His access to HIV care provided a source of information and support since he expected to be at high risk for COVID-19 disease based on his medical history.

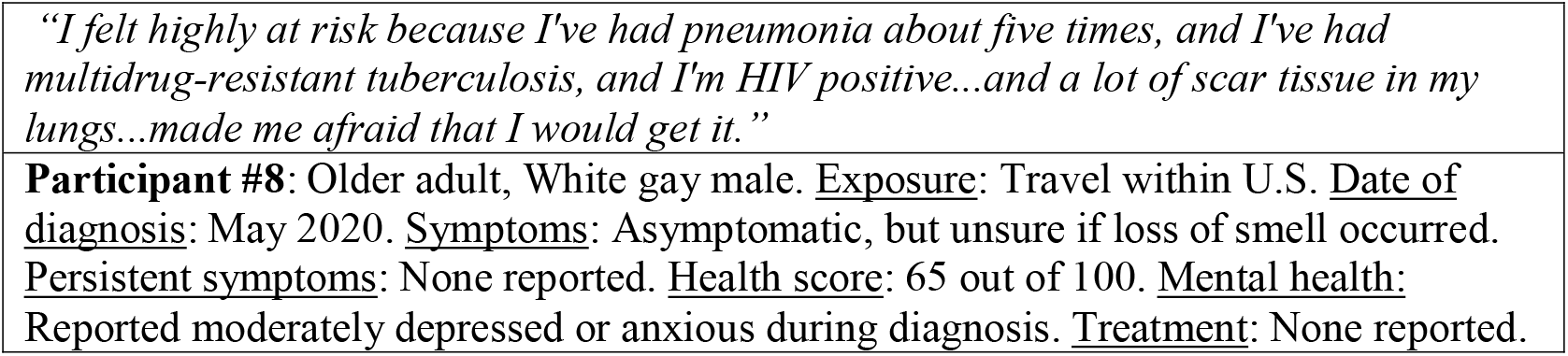

Another factor in their recovery was how participants could actively benefit from their health information-seeking behavior by having access to medical care and providers.

*“…My doctor is an infectious disease specialist, and I asked her about it*.*…I asked my nurse about it, who is in the same clinic as my doctor. And also news and media outlets. I just tried my best to look for the facts*.*”*

Similarly, participant #24, a Spanish-speaking woman and caregiver to her two children who had serious medical conditions, and whose health score was 70/100 before her diagnosis, described the support she had by having access to a medical team after testing positive for SARS-CoV-2 prior to surgery. The following quote also highlights how she actively sought COVID-related health information.

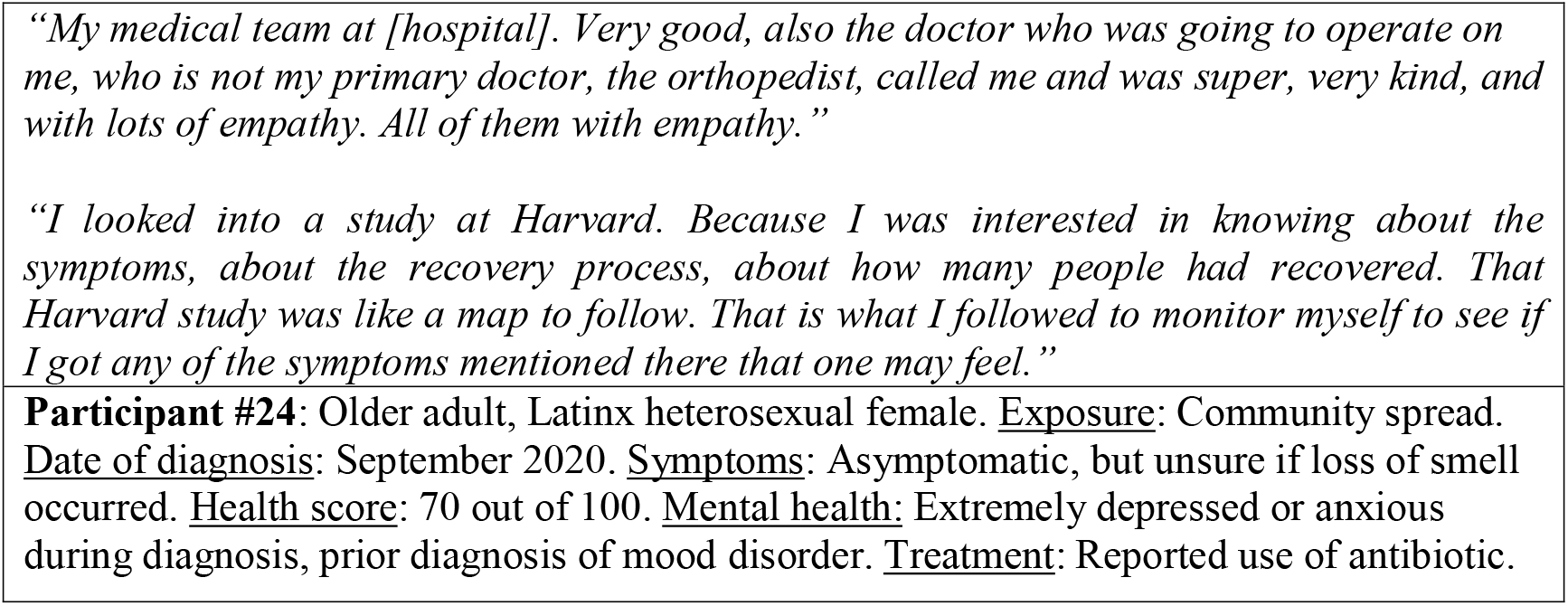

After receiving her diagnosis, participant #24 explained that her primary care provider also offered her support as she anticipated the onset of illness symptoms.

*“I stayed in touch with my primary doctor, he was a great support. He told me that if I had trouble breathing or if I had a high fever. But no, I monitored myself and, thank God, I didn’t have any symptoms. If truth be told it was only boredom*.*”*

For health information-seeking behavior to benefit a person, it must result in a change in outcomes, such as new knowledge gained or receipt of support, both of which were present in the narratives of participants #8 and #24. For example, when participant #8 stated he sent multiple e-mails to his HIV medical provider to follow up on his original inquiry, he was able to receive moral support. His information seeking also led to his awareness of ongoing discussions of whether antiretroviral therapy was offering protection to people living with HIV.

*“She [HIV provider] didn’t respond because when I did finally catch up to her, I realized how busy she was, because that’s what she does for a living. She’s an infectious disease doctor…I think the only support I needed from her was just moral support…And the nurse that I have*…*she’s really good, and she reached out to me…they called about five or six times after that to ask if I needed anything, if I needed food, if I needed this, that, the other thing*.*”*

*“I heard people saying people who are on lots of antivirals, like myself, weren’t getting it as readily, but I don’t know the facts behind that*…*”*

Despite the reassurances that participant #8 received about his risk for severe COVID-19 symptoms because he was living with HIV, he was also aware of his risk for severe COVID-19 due to his diabetes.

*“Well, just the fear…my immune system has been intact, so I assumed that I would be okay. But I believe I was actually more concerned with diabetes, although I wasn’t informed about how COVID affects diabetes. And I still don’t know to this day. And I was just more worried about that than I was about my HIV*.*”*

In documenting the “recovery” process from asymptomatic infection, the importance of social support was also evident throughout the participants’ experiences. As participant #8 stated:

*“…I actually didn’t have time to worry because they called me the next day. I had heard that the test results come back in like three to five days, and when they called me the very next day, I was kind of really shocked. I was like, ‘What do I do now? I didn’t know what to do. Then I called my partner immediately, and I said, ‘I got a positive result*.*’ And he said, ‘Well we’re going to quarantine then, for two weeks,’ which we did. Nobody came and nobody left*.*”*

Participant #24 had a partner whose occupation involved cleaning, and thus, he cared for her and the household during her isolation period while recovering from COVID. She thought that the cleanliness may have helped in her process of recovery.

*“He always came, he cleaned me, bathed me. He disinfected everything. When we had COVID, I don’t know if that may have been our success, maybe it is just my theory, because just by chance my husband works in the cleaning business*.*”*

Most importantly, for both participants, there was an acute experience of psychological distress during their diagnosis. Participant #24 stated that while she had no physical symptoms from COVID, she was affected emotionally.

*I didn’t have any symptoms, but it affected me psychologically, because knowing that you have coronavirus impacts you emotionally and one lives with the uncertainty of not knowing what is going to happen. You ask yourself: How is it going to hit me? And if I die? And if I stop breathing? I am a very sociable person. And isolating yourself, even if you know it is for a good cause, makes you depressed. I even got to cry and that ‘Ok. If I have to die, I have to die*.*’*

Participant #8 expressed concern about his asymptomatic infection considering other serious medical conditions he had experienced before.

*“I’m not a fearful person. I’ve almost died a couple of times, so nothing really bothers me or frightens me. But this is a little scary…I was afraid. I was concerned with getting it because I’ve had all those infections*.*”*

After receiving support from their medical teams, but still maintaining a sense of distress over the uncertainty of the disease course, experiences of perceived stigma due to COVID-19 were present among many participants, as this quote from participant #24 shows.

*I still have to teach people. ‘Look, here it is, it is evidence. I have two not just one, negative tests. Because even afterwards they do not want to get together with you, because they say: ‘And the people who have had it are not contagious? ‘No, if I am already negative. How can I give you something that I don’t have?’ That is common sense. I don’t need to be a scientist*.

### Symptomatic Infections

Participants’ symptomatic experiences with COVID-19 varied by clinical features, ranging from mild symptoms to symptoms of acute respiratory distress syndrome. Many reported persistent symptoms at the time of study enrollment. Several participants believed their initial COVID-19 symptom onset could be explained by another cause. In some cases, they attributed initial symptoms to seasonal allergies or influenza. For instance, participant #15 originally confused her symptoms with allergies. Her health score during her COVID-19 illness was 60/100. She shared:

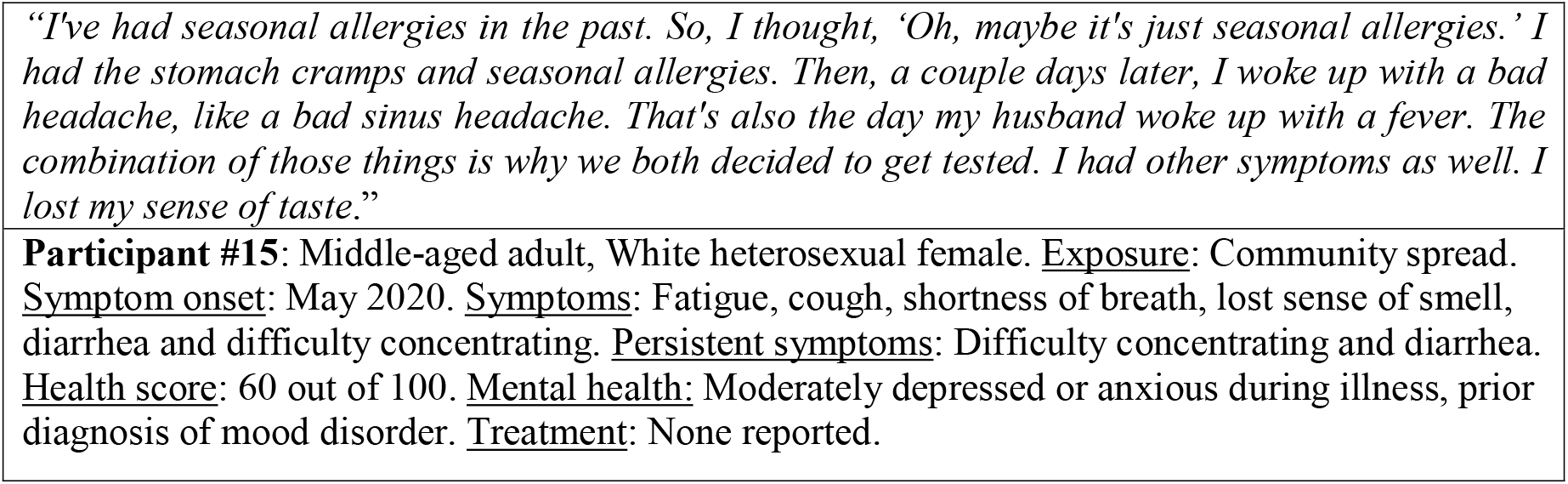

Both participants #17, a middle-aged female (did not report a health score) who required hospitalization, and participant #21, who reported a health score of 60/100, took their initial symptoms for something else; menopause and a common cold. Although they reported similar symptoms, their characterization of these symptoms was different.

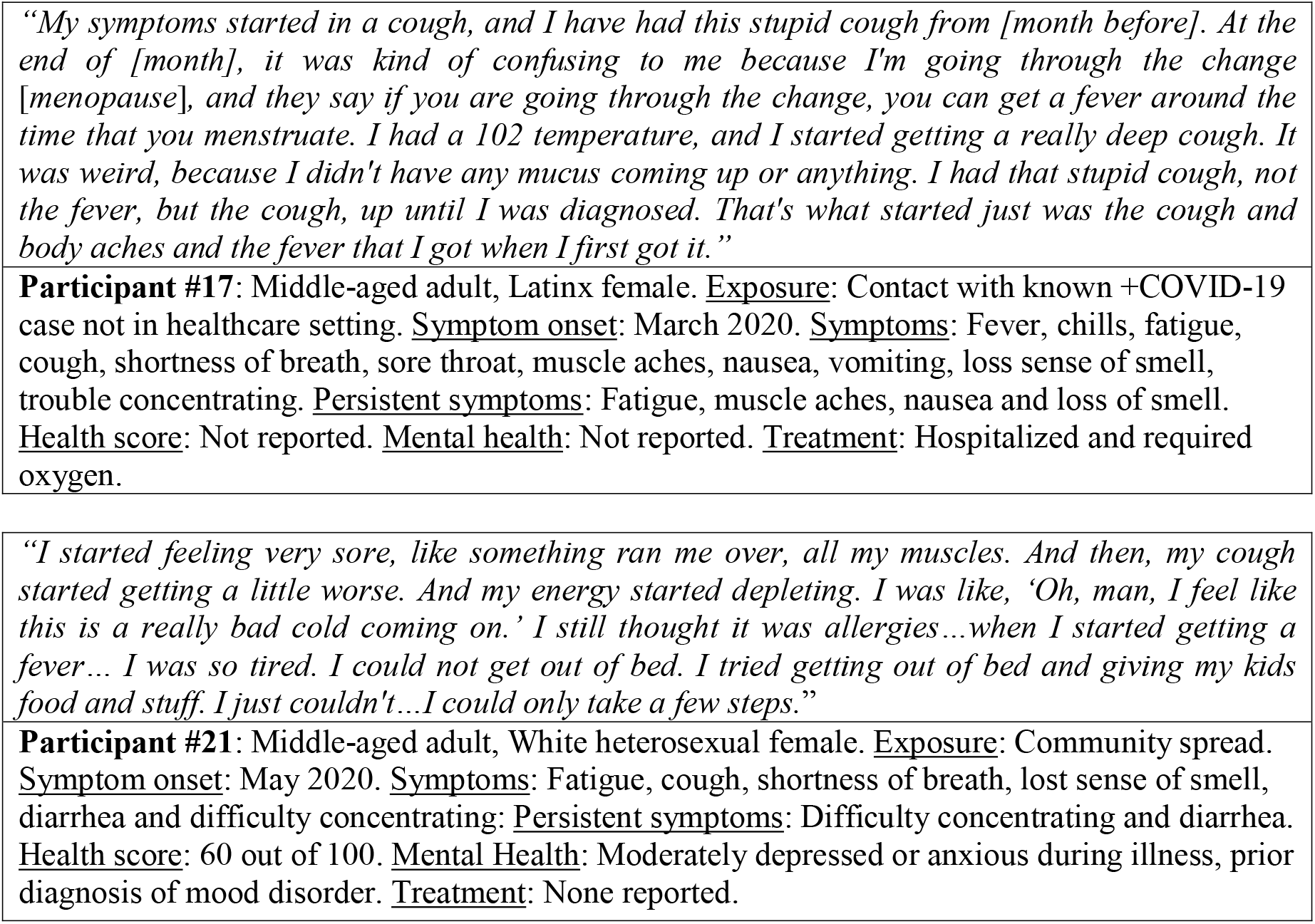

Two additional participants, #6 and #7, reported similar symptoms to those mentioned directly above, but also suffered from shortness of breath, loss of taste and smell, and extreme exhaustion. Participants #6 and #7 provide insights into the rapid onset of severe illness and a feeling of getting “hit like a train” all at once unexpectedly.

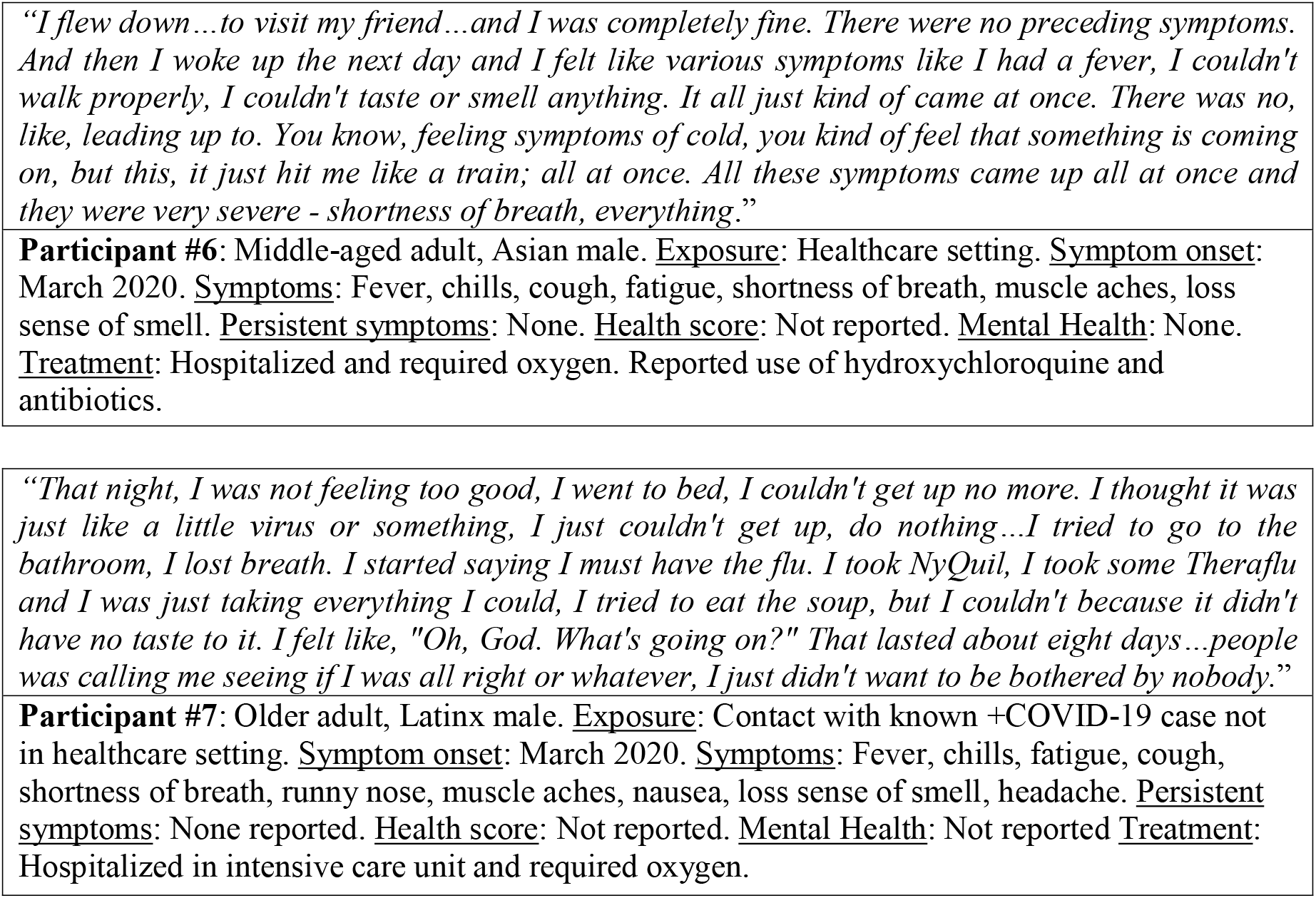

The five narratives above (#15, #17, #21, #6 and #7) showed that while similar symptoms were reported, their experiences with COVID-19 were heterogeneous. Some participants’ symptoms persisted longer than expected, even as assessed by their healthcare providers. Participant #4 shared that after 14 days with a fever, his symptoms intensified. He also emphasized deficits in memories and awareness of the severity of his symptoms, and the importance of the support he received from friends.

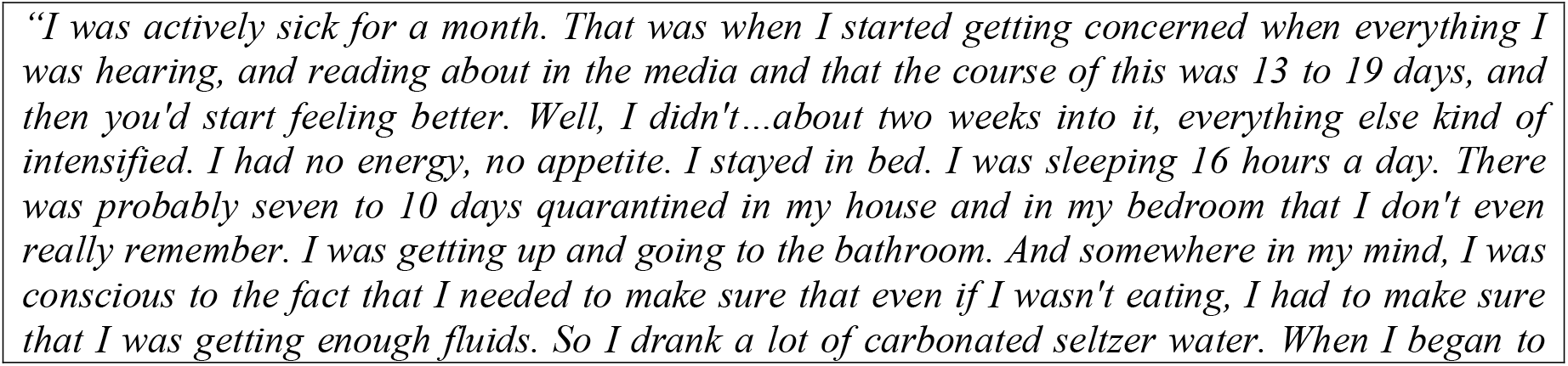

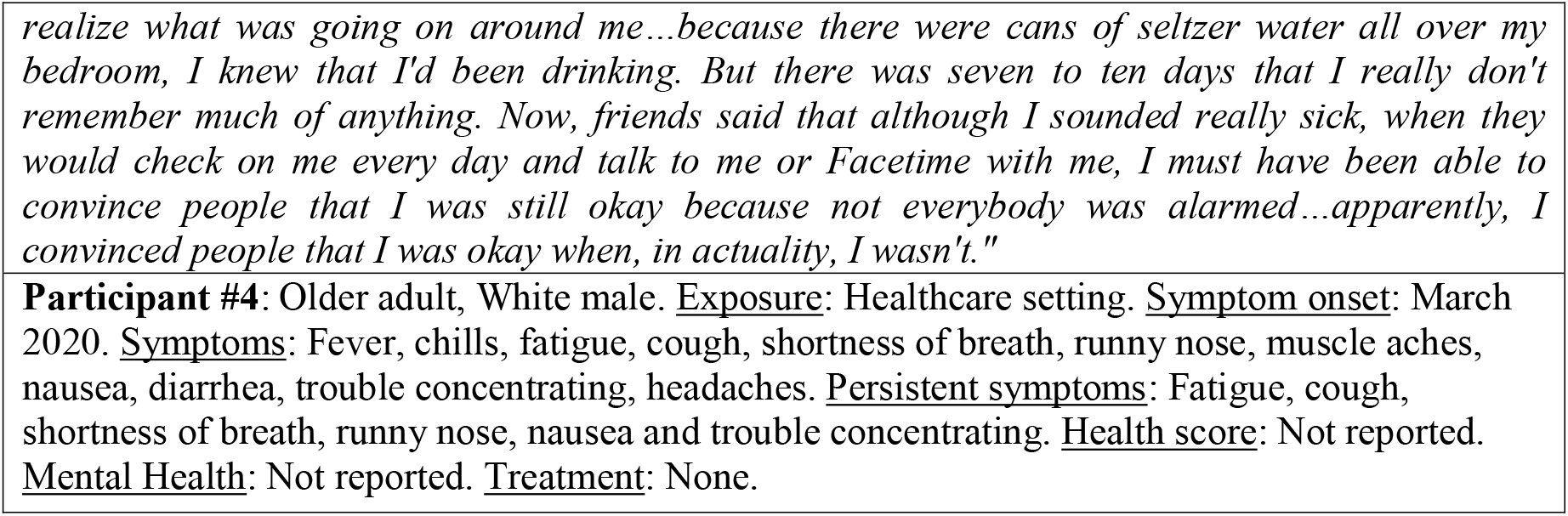

Similarly, participant #14 shared that her symptoms were worse later in the disease course. Her symptoms worsened on day 19, and she was not fully aware of what was happening.

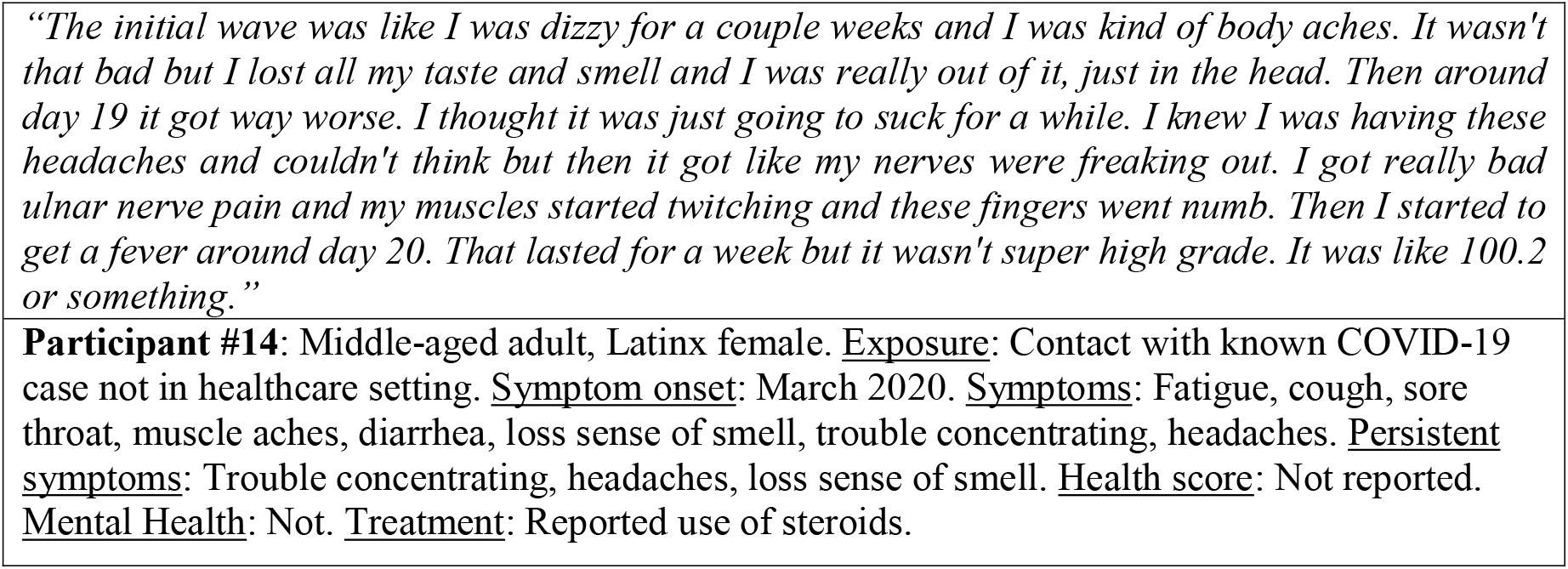

The experiences of participants #4 and #14 highlight the severity and burden of managing persistent COVID-19 symptoms. While the clinical evaluation data show overlapping symptoms across all the participants quoted above, their experiences were variable but intense. One common thread was confusion and uncertainty – either from processing the rapid onset of severe symptoms with their initial diagnosis or concerns regarding cognitive impairment following recovery from the acute process. While we did not capture full information on the burden of managing symptoms that persisted at the time of the interview, we characterize the initial experience as very burdensome, including ongoing uncertainty among several participants. In the next section, we characterize the experiences of people who accessed medical care for COVID-19.

### Experiences with Hospitalizations and Advanced Medical Attention

Nine of the participants we interviewed accessed emergency room services, had prolonged hospitalizations or, even, intubation. Participant #22, a Spanish-speaking male, who was hospitalized for a month shared that he was unconscious for over a week and had no recollection of what happened during that period.

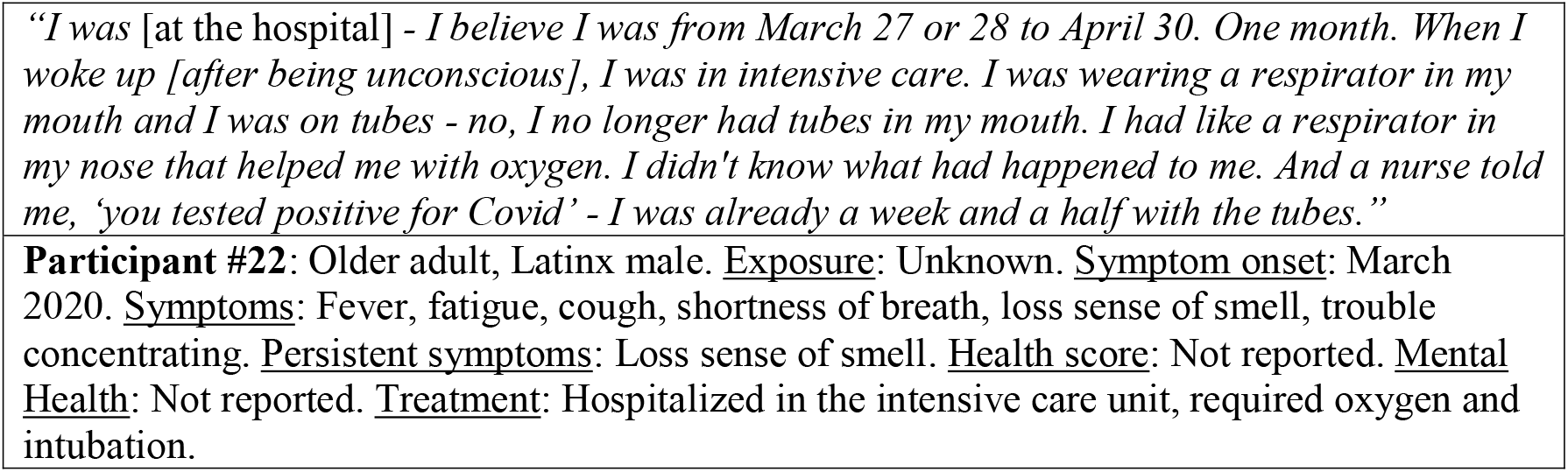

Another participant spent 35 days at the hospital, 20 of which were in the intensive care unit. He required intubation and recalled hallucinatory-like experiences (unclear if auditory or visual) during that time. He also shared not being able to communicate by writing.

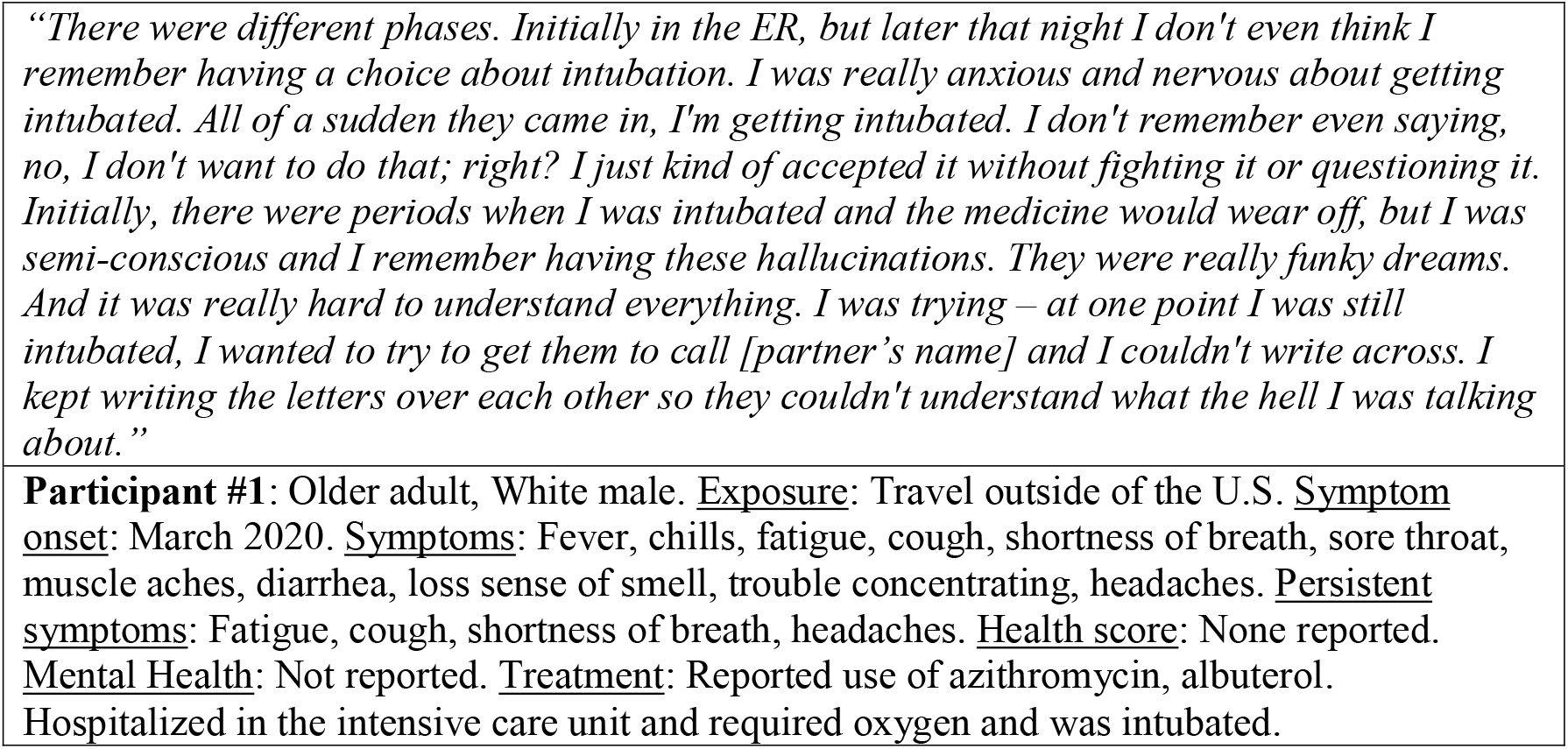

Participant #21 (described further above) was also hospitalized and suffered memory loss. In her case, she was taken to the hospital after losing consciousness during her COVID-19 illness, suffering a broken nose and head injury.

*“I decided to work a little bit because I felt like, oh, I have a great energy today. That’s how it happened in the mornings. I had amazing energy. But then, an hour into it, or maybe two hours, I was completely done. And I was feeling miserable. So, that day, I tried to work from home. And after an hour, I said, ‘No. I am feeling miserable. I’m going to go lay down and sleep*.*’ But I woke up*… *I was drenched in sweat, and my stomach was very upset. And I need to go to the bathroom, and I need to throw up. And I came back to turn on the fan. And I don’t remember seeing anything after that. I might have run back to the bathroom. And then, I remember someone screaming to see if I was okay. So, I ended up fainting because I was very diaphoretic and pale. And my blood pressure apparently tanked. So, I broke my nose and split my head open. And I had to be taken to the hospital*.*”*

Common experiences among participants hospitalized were clear gaps in memory, confusion, and other neurological features, including persistent symptoms of trouble concentrating, headaches, and loss of sense of smell. The consequences of these experiences remain unknown, but warrant further investigation to determine the management and recovery process of participants with severe acute disease and persistent symptoms.

### COVID-19 Anxiety, Frustration and Stress

COVID-19 affected the mental health of all participants, including participants with asymptomatic infection. This was attributed by both asymptomatic and symptomatic participants to either the uncertainty behind their diagnosis, symptom course, the initial severity of symptoms for some, the fear of dying, and concern for the health of family and friends. In some instances, predisposition to experience psychological distress were catalyzed by COVID-19. During the interviews, we asked participants what went through their mind while awaiting test results.

Participant #24 (asymptomatic case described above) shared her feelings of shock:

*“I didn’t have any symptoms but it affected me psychologically, because knowing that you have coronavirus impacts you emotionally and one lives with the uncertainty of not knowing what is going to happen. You ask yourself: How is it going to hit me? And if I die? And if I stop breathing?*…*And isolating yourself, even if you know it is for a good cause, makes you depressed. I even got to cry and that ‘Ok. If I have to die, I have to die*.*’”*

Participant #9 expressed the fear of an unknown disease course following diagnosis, but also noted his relief from expected immunity after recovery.

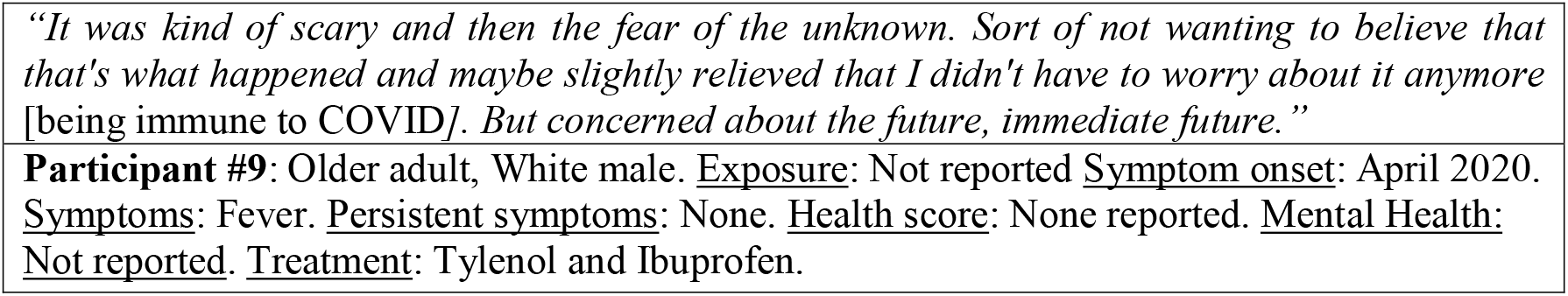

Participant #5 shared his struggle with depression, compounded by the process of social isolation due to his diagnosis. His fear of dying alone led to a psychiatric emergency requiring hospitalization.

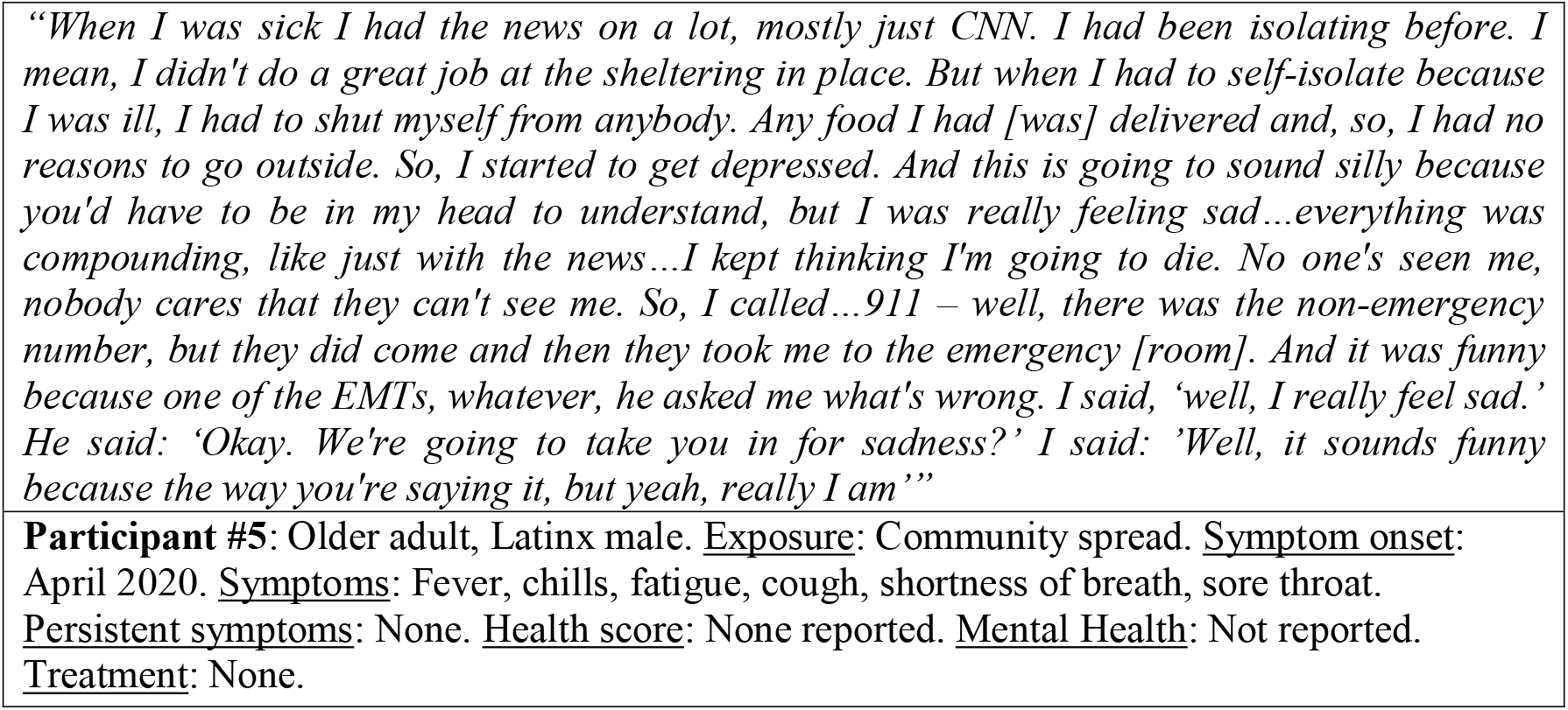

Participant #6 (described above) described feeling anxiety and frustration over his recovery process:

*“At that point I was, like, I probably have it. But as far as recovering, that’s what I was more worried about; like, when is this going to be over? Like I don’t feel I’m getting better. If anything, I’m declining. So, it was more of like a feeling of anxiousness and, like, frustration*.*”*

Levels of stress, anxiety and frustration among participants could be directly related to the fear of dying or uncertainty pertaining to the health consequences of COVID-19. Participant #14 (described above) shared that he could identify the symptoms and knew it was COVID-19, and mainly worried about the possible progression of his situation:

*“I knew that I had it probably. I guess, I don’t know, it’s just like, ‘Okay*.*’ At that time, I would have been a lot scared now because we know how crazy it can be and then people fall off a cliff. We didn’t know any of that back then, so I was like, ‘You know, as long as I’m not dying then I guess I’m not dying’…I was worried because I had done enough research to understand that the receptors where your taste, where your smell, are very close to the brain and the central nervous system so it’s like, ‘Oh God, what if it gets into my brain. We don’t know anything about this*.*’”*

### HIV and Covid-19

As the parent study was housed within an HIV care and research center, we were able to recruit people into this study who were living with HIV and recovered from COVID-19. Most of the participants living with HIV were very attuned to early concerns of being at higher risk for severe COVID-19, although the fear of having more severe outcomes in this population attenuated over time with more data. Overall, participants noted that the fact that they were living with a co-infection of HIV facilitated them seeking information, staying vigilant, and following public health guidelines.

Participant #23 stated feeling very fortunate, despite “my condition” [HIV], having had a few critical days when he felt “deep anguish”, and compared himself to other people of different ages he knew or had heard of that died of COVID-19, including two family members in Mexico.

**Participant #23**: Older adult, Latinx gay male. <u>Exposure</u>: Community spread. <u>Symptom onset</u>: July 2020. <u>Symptoms</u>: Fever, chills, fatigue, cough, shortness of breath, sore throat, muscle aches, loss sense of smell, headaches, irritability. <u>Persistent symptoms</u>: Headache. <u>Health score</u>: 50 out of 100. <u>Mental Health</u>: Moderately anxious and depression during illness. <u>Treatment</u>: None reported.

Participant #5 (described above) felt at higher risk because of his HIV status and additional comorbidities:

*“I kept thinking I was one of the ones that if you got it, it’s really not going to look good for you. I’ve had two heart attacks. I have HIV…viral load is not so great. Yeah, just didn’t think it would bode well for me to have the COVID-19*.*”*

He explained that usually he struggled with adherence to his HIV treatment but that while recovering from COVID-19, he tried to regularly take his medications “with a purpose”, having heard on the news that antiretrovirals may have a protective effect.

*“I’m not really good at taking my pills, so that’s a separate problem, but I took them as much as I would probably have ever taken them. I started to hear on the news something about the HIV medications or – it’s kind of – it was good to – you could combat the virus. I don’t know. There’s something in there, some component. So I thought: Oh, God, you better really take your meds’”*

In most cases, participants stated that their HIV care providers called them regularly to provide support and reassurance. Participant #19 (described further above, Spanish-speaking Latinx gay male) explained that his doctor had reassured him that his HIV would not affect his recovery process.

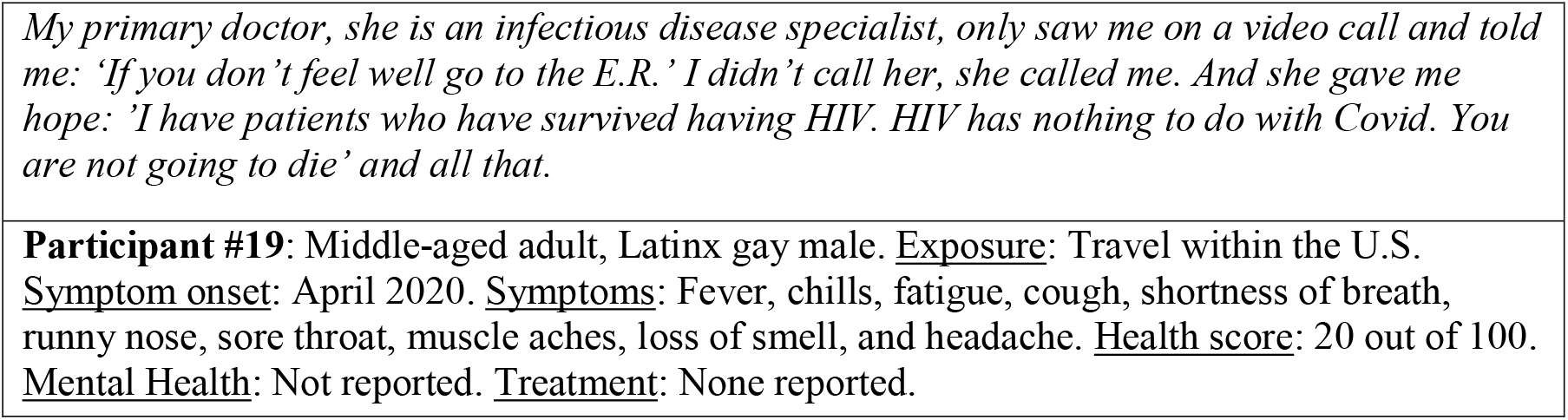

Participant # 10 explained that he initially had nose bleeds and headaches, but not having seen those in the list of COVID-19 symptoms, he did not call his HIV provider because he was afraid he would be told he had COVID-19. However, he acknowledged that he could have addressed this concern if he had contacted his HIV care provider.

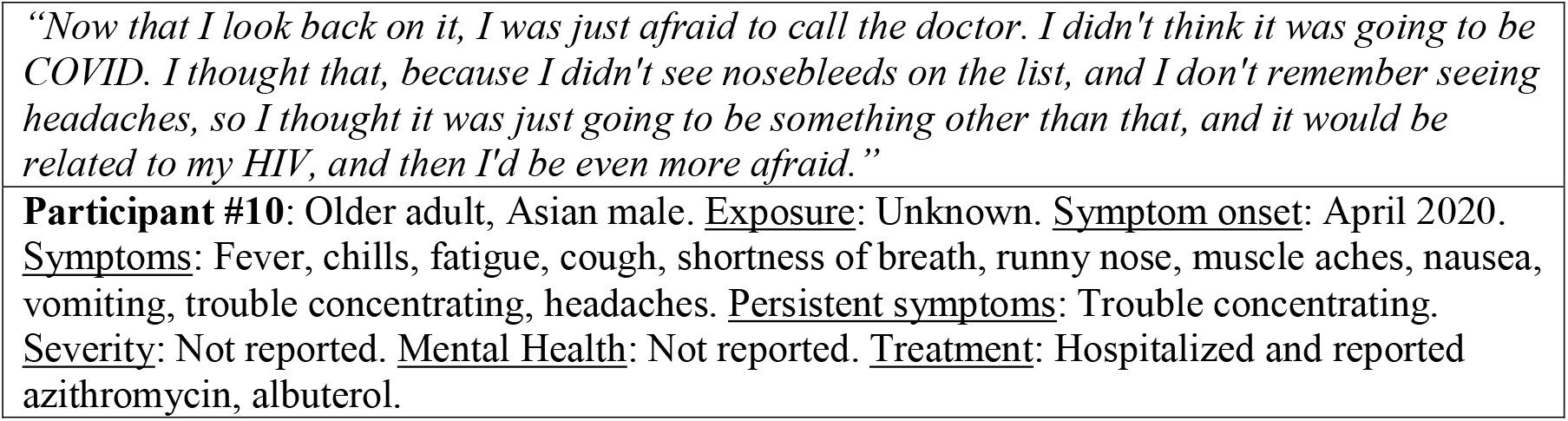

Participant #10 expressed the feeling of being afraid of dying from COVID-19, but the support from his neighbors and friends, his family to some extent, and knowing that a person delivering food from Meals on Wheels (non-profit organization providing food assistance) was on the other side of his closed door, sustained him during his recovery process. He explained that his family was concerned for him but qualified his family support by saying they did not know how to deal with a crisis and had asked him whether he had made “arrangements” in preparation for his death. He noted that he only recently disclosed his HIV status to his family, which may have influenced their reaction.

*“One of the things about when you’re sick, after the diagnosis and you’re in quarantine, you kind of start to give up. I started to think that, maybe it’s just faster to just take care of it, to end it. Because, all I’m doing is sitting here and being sick. You’re just thinking you’re going to die. There’s nothing saying that you’re going to live, and I have HIV, I’m in a high risk category. Nothing was giving me any sense that I was going to live. So, why would I want to go through this, and just continue to keep going through this? That’s why it was important for me that, when the Meals on Wheels people came, that little bit of contact with the real outside world and not through Zoom, it helped a lot. I’d wait until they’d come to deliver the food. All they’d say is, “Your food is here,” and I’d say, “Thank you,” but I’d look forward to it*.*”*

Like all participants in our study, participant #10 was trying to reconcile uncertainty around immunity and risk of re-infection, which was yet to be determined in the scientific literature at that time. Like other participants, he was actively seeking health information through a local and well-known HIV-related institution in San Francisco. Further, he deliberated over whom to tell he had had COVID-19 in anticipation of community stigma. He emphasized the importance of the lessons learned through time from the HIV epidemic, and wanted to acquire the skills to manage his fear and stigma related to COVID-19 and how to reinsert himself back to social life:

“*What I need to understand is how people with HIV assimilated back into the community. In the early days, how did they do that? I don’t know how to do that, and that’s the thing I’m most afraid of, that I won’t be able to*… *I’m afraid that I’m going to isolate myself further, because I don’t know how to join back in, and how to deal with the stigma. That I’m going to get more of it. I’m a realistic person. When people are afraid, if people are afraid, I’m going to get more of it. I need to know how to better handle that. My plan was to go to the SF AIDS Foundation, to see if they have some ideas and thoughts on how to re-assimilate, when you’re faced with the fear of others*.*”*

Importantly, a participant who was not living with HIV compared the experience with COVID-19 to the initial reactions during the HIV epidemic in the 80s. Participant #14 (described above) stated:

“*I would just say it’s very scary. It feels like, it must feel like when people were getting diagnosed with HIV early on. We didn’t know what it was and it’s very unclear what it means for you*.*”*

Participant #11 described the lack of knowledge of the possible impact of a novel virus in society in general. Similarly, another participant talked about the uncertainty and fear associated to a new virus but added that the COVID-19 and HIV epidemics are natural phenomena.

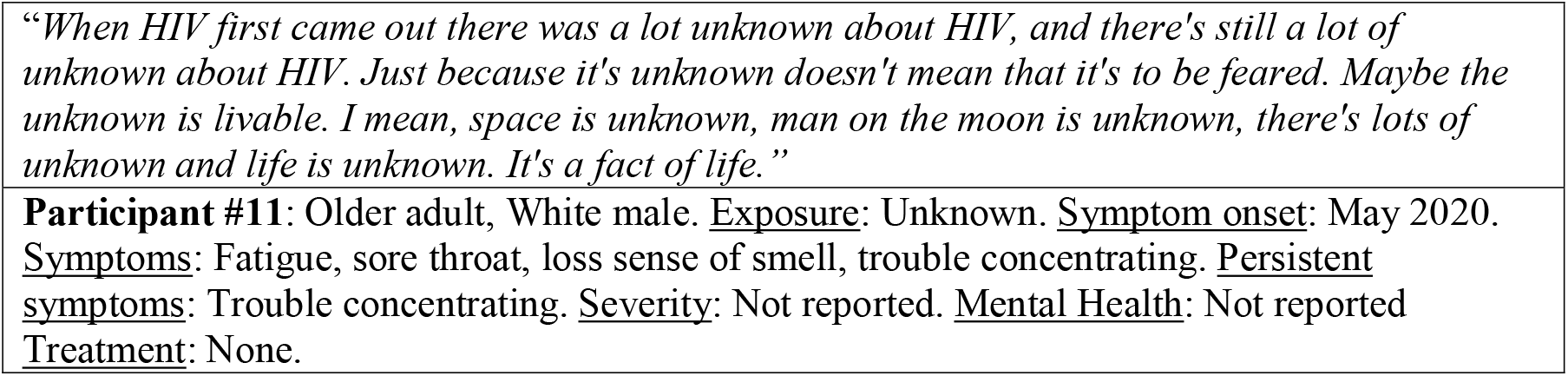

In summary, participants who had been living with HIV had a heightened awareness of the risks for severe COVID-19 disease, drew lessons for coping from the HIV epidemic, and also displayed high levels of health information-seeking behavior, in part due to their engagement in HIV care. Critical lessons can be learned from the participants in this study related to management of persistent symptoms with COVID-19, ensuring easy access to health information and wrap-around service support, including mental health support.

## DISCUSSION

Our qualitative data from English- and Spanish-speaking individuals with a past diagnosis of COVID-19 across a variety of disease presentations illustrate the challenges of understanding and evaluating the variability in the illness and recovery experience with COVID-19. Overall, three findings emerged that were central to the acute experience with COVID-19 that we believe may be useful for research on post-acute sequelae of COVID-19. First, all individuals had to manage pervasive uncertainty in multiple domains, including processing the possible consequences of asymptomatic infection; uncertainty regarding the course following symptomatic infection in terms of cognitive function and memory loss; and the ongoing recovery process, especially among those requiring hospitalization. Second, given that COVID-19 is a novel disease, we found high levels of health information-seeking behavior (e.g., calling medical providers, sources of trusted information). Importantly, information-seeking behavior served as a proactive coping strategy for managing the unpredictability of the illness at the time and facilitated by access to medical care. The resulting information-sharing participants received from medical staff and providers led to receipt of instrumental, moral, and social support. Lastly, despite different symptom profiles and severities, all participants shared similar levels of psychological distress, likely through the underlying uncertainty they were attempting to cope with both related to COVID-19 and related scenarios (e.g., implications for other medical conditions, concerns for family, mental health).

### Implications

Recently established cohort studies show that common symptoms among people with post-acute sequelae from COVID-19 are frequently non-specific: fatigue, myalgias-arthralgias, and neurological (i.e., headaches, “brain fog”),^7,21, 22^ suggesting that managing patient uncertainty will be a part of the overall coping process and management. Our data shed light on the intensity of the illness experience, even across initial symptom strata, and suggest that psychological adjustment will be key in recovery. While neurocognitive post-acute sequelae is an emerging and less understood area,^23^ there may be implications for these sequelae exacerbating psychological distress resembling post-traumatic stress (i.e., arousal, worry, intrusive thoughts) given many participants had near-death experiences.^24-27^ But importantly, in identifying frameworks to help understand the experience of a new disease with a high degree of uncertainty, we found lessons can be drawn from management of chronic diseases (e.g., pulmonary disease, liver disease, and HIV).

Our data regarding how participants processed and coped with their illness showed similarities to the dimensions of the healthcare empowerment model, originating out of work in HIV, cancer, and cardiovascular disease. This model outlines that coping with a novel disease such as COVID-19 will require navigating a level of uncertainty, which can be managed by supporting actions to stay informed and engaged with a treatment plan and team.^13,28^ Other research from pulmonology and gastroenterology,^29,30^ including interventions and qualitative research, points to use of cognitive and behavioral strategies to manage psychological distress over the unpredictability of illness and prognoses, and recommends communication with healthcare providers about their own clinical uncertainties. As participants’ experiences highlighted, there will be a need to provide information and continuous support to persons with post-acute sequelae to ensure they feel secure along the path to recovery.

## Limitations

Our study has several limitations. First, it was completed amid the first two waves of COVID-19 cases in the United States, and new information and details about the virus, symptoms, treatments and sequelae were evolving daily. Second, our participants were engaged in the start of a newly developed cohort study; these engaged participants may differ from other populations of individuals who have recovered from COVID-19. Because the study took place in an infectious disease (including HIV) care and research center in San Francisco, which took aggressive measures to shelter-in-place earlier than other major cities, and many participants were already linked to the medical setting formally, there are likely differences in experiences compared to persons with limited access to medical care. And while we presented clinical evaluation data alongside selected quotes, these data were not used or reviewed by interviewers at the time of the interviews and thus, we are limited in our ability to integrate these data with our qualitative data. However, given the patterns that emerged across all narratives, we have no reason to believe that the results depend on other characteristics of the participants or context.

## Conclusion

We conducted a qualitative study of individuals who had recovered or were recovering from COVID-19 with wide-ranging experiences. We were able to leverage personal narratives of experiences with asymptomatic infection, symptomatic infection, including hospitalization, and HIV co-infection, to identify patterns in how the disease was experienced and gain insights into the recovery process. Psychological distress was elevated regardless of symptomatology. Information-seeking behavior, health empowerment, and social support were enabling factors toward recovery for our participants. These data may help shed light on the management of emerging post-acute sequelae of COVID-19, including addressing issues with disease uncertainty, mental health, and persistent symptoms.

## Data Availability

The data are available with a formal request to the principal investigative team and with a data sharing plan to protect against breaches of confidentiality.

https://www.liincstudy.org/

## Acknowledgements

We are grateful to the LIINC study participants and to the clinical staff who provided care to these individuals during their acute illness period. We acknowledge LIINC clinical study team members: Tamara Abualhsan, Mireya Arreguin, Jennifer Bautista, Monika Deswal, Heather Hartig, Marian Kerbleski, Sadie Munter, Fatima Ticas, and Meghann Williams. We acknowledge LIINC leadership team members Bryan Greenhouse, Isabel Rodriguez-Barraquer, and Rachel Rutishauser.

